# Beyond conventional statistics: Genomic Informational Field Theory (GIFT) identifies sex-specific herpes virus associations in multiple sclerosis

**DOI:** 10.64898/2026.08.04.26359688

**Authors:** Naqib Ahmed, Peter Maple, Radu Tanesescu, Lucia Giorgi, Paola Valentino, Alessia Di Sapio, Bruno Gran, Cyril Rauch, Karim L. Kreft

**Author notes:** Deceased (Peter Maple). **Corresponding authors:** Dr. Karim L. Kreft and Dr. Cyril Rauch University of Nottingham, Derby Road, NG7 2UH, Nottingham, UK.

## Abstract

**Background:** Detecting higher order relationships in datasets of complex traits, such as multiple sclerosis (MS), has been challenging. Conventional statistics largely rely on comparing averages across groups and thereby discard important information on the underlying distribution of datapoints. The Genomic Information Field Theory (GIFT) overcomes this limitation by ranking individuals based on linear measures, for example immunoglobulin titres. The exact role of humoral immune responses against several human herpes viruses in a sex-dependent manner in MS is currently unknown.

**Materials and methods:** We compared the performance of GIFT with conventional statistical frameworks to detect differences in the humoral immune response against 4 highly prevalent herpes viruses linked to an individual’s susceptibility to develop MS in 200 MS patients and 137 healthy controls.

**Results:** GIFT validated the well-known association that the Epstein Barr Virus (EBV) protein EBNA1 is strongly linked to MS susceptibility in both sexes. In contrast to conventional statistics, GIFT also identified association between herpes simplex virus, varicella zoster virus and the EBV VCA protein and female susceptibility to develop MS, whereas male MS susceptibility was only linked to CMV immunoglobulin levels. None of these associations was observed using conventional statistical tools.

**Conclusion and discussion:** We here show for the first time that GIFT is able to detect novel associations in human immunoglobulin data linked to MS susceptibility, which remained undetected by conventional statistical frameworks. This shows the power of GIFT to detect complex phenotype-trait associations and underlying subgroups within populations.

## Introduction

Multiple sclerosis (MS) has considerable sex-specific effects. It affects woman approximately three times more often than males, and the sex-ratio is increasing over time, mainly driven by an increase in MS incidence in woman [1]. Females are generally younger at onset of symptoms [2]. However, males have a shorter time to secondary progressive MS and are relatively younger when transiting to progressive MS [3], indicative of accelerated neurodegeneration. Moreover, the development of brain atrophy and cognitive impairment is significant more severe in males [4]. However, the underlying pathophysiological mechanisms driving the sex differences is currently poorly understood. Risk factors increasing the susceptibility to develop MS are both genetic variants [5] and environmental factors, most notably the herpes virus Epstein Barr Virus (EBV) [6]. Virtually all MS patients have been exposed to EBV [7]. EBV infection precedes an MS diagnosis [8] and studies in familial MS provide support for EBV-host genetic interactions [9]. Recently, increasing interest in the role of other herpes viruses in MS has emerged, and a meta-analysed found that MS susceptibility is also linked to varicella zoster (VZV) and human herpes virus 6 (HHV-6), besides the well-established EBV association [10].

Viral infections also have strong sex-specific effects, for example cytomegalovirus (CMV) has a higher prevalence in females, whereas EBV-associated diseases are more common in males [11]. To complicate matters further, herpes simplex virus 2 (HSV-2) have both sex- and age-specific effects [12]. Taken together, the complex sex- and age-specific effects of herpes viruses and their potential effect on MS susceptibility requires sophisticated analyses. Conventional statistical frameworks are relatively limited in identifying potential complex associations particularly through their requirement of very large sample sizes to detect small subgroups and their reliance on comparing averages across groups rather than utilising the full distribution of datapoints.

By revisiting the mathematical foundations of conventional statistics, we identified key limitations concerning their ability to extract information from datasets and in particular the fact that a substantial amount of information is lost during analyses [13–17]. Conventional statistics use frequency- or count-based visualizations, such as bar charts or histograms, which group raw data into discrete bins. While these discrete representations provide the foundation for calculating key population-level statistical summaries (e.g., means, variances, p-values etc.), which in turn underpin dataset-level inferences, they do so at a cost. By transforming raw data into aggregated representations, the process necessarily sacrifices information about the individual datapoints. To illustrate this point, consider a dataset where raw data measurements are captured with high precision, rendering each data point unique, a scenario increasingly frequent with the advent of advanced measurement technologies. Once these data are grouped into bins to construct frequency- or count-based plots, the individuality of each measurement is effectively erased, as all individual datapoints within a bin become indistinguishable. As a result, a substantial amount of information is lost through this methodology. To overcome this limitation and recuperate the information lost by conventional statistics, we have devised a new methodology a.k.a. genomic informational field theory (GIFT) inspired by field theory in physics and information theory [15–17]. Validated in the field of quantitative genetics, i.e. genotype-phenotype mapping [13,14], our aim is now to generalize the application of this method to other fields of research, e.g. quantitative protein measures such as humoral immune responses against herpes viruses in MS.

We hypothesize that herpes viruses have sex-specific effect on MS susceptibility and applied GIFT to a cohort of 200 deeply-phenotyped MS patients and 137 age matched healthy controls and assessed the effect of herpes viruses on susceptibility to develop MS.

## Materials and methods

### Clinical data

200 multiple sclerosis patients from the Multiple Sclerosis Regional Referral Centre Biobank (CRESM Biobank) at the University Hospital San Luigi Gonzaga, Orbassano, Italy, were followed prospectively and blood samples were collected prior to start of disease modifying treatments. Serum samples were processed within 2 hours and stored at -80°C. 137 healthy control samples were collected using the same protocol. Details about sample collection and storage have been published elsewhere [18].

### ELISA for immunoglobulin levels against herpes viruses

Immunoglobulin G (IgG) levels against EBV epitopes (Viral Capsule Antigen, VCA; Epstein Barr Nuclear Antigen-1, EBNA-1), varicella zoster (VZV), herpes simplex 1 and 2 (HSV1/2) and cytomegaly virus (CMV) were measured using commercial assays. ELISA plates were coated with 100 ml calibrators and included positive and negative control sera (Institut Virion/Serion, Würzburg, Germany). Serum samples were diluted 1/100 in assay buffer, incubates for 1 hour at 37°C, subsequently the plates were washed and incubated with anti-human IgG conjungate for 30 minutes and washed again. Para-nitrophenylphosphate substrate was added, and the plates were subsequently incubated for a further 30 minutes. The reaction was stopped and optical densities (OD) read at 405 nm (reference wavelength 650 nm, BioRad Benchmark Plus microplate spectrophotometer). Quantitative antibody levels were determined by interpolation of OD values using manufacturer-supplied standard curves. If a sample was above or below the linear range of the assay, we assigned a value one step above or below resp. the actual value. Details have been published elsewhere [18]. We tested ELISA coefficient of variation, and this was routinely <10%.

### Ethical approval

This study was approved by the Ethical Committee of San Luigi Gonzaga University Hospital (CRESM Biobank approval, ref 18390/2019) and Nottingham Research Ethics Committee 2 (Ref. 08H0408-167) and samples were handled in line with the UK Human Tissue Authority guidelines. All patients and controls provided written informed consent prior to inclusion.

### GIFT paths

Standard GIFT paths are determined by ordering the individuals by viral titre and then summing their microstates (e.g. MS versus control) cumulatively along the ordered sequence. MS patients and healthy controls were assigned microstates +1 and -1 respectively.

To account for non-unique viral titres, ‘smoothed microstates’ are calculated by taking the mean of the microstates of individuals with the same viral titres. Taking the cumulative sum of these smoothed microstates then gives the most informative path that can be formed from the data.

### Statistical Analysis

While a useful criterion to find a p-value already exists, the presence of non-unique viral titres means that the formulation is not accurate in this case, and we must use a revised method. In this paper, the test statistic used is the range between the maximal and minimal value $\theta_max - \theta_min$. P-values are obtained by comparing the observed ranges with paths simulated under the null hypothesis. We simulate such paths by randomly permutating 10,000 times the microstates among the individuals, effectively removing the relationship between the microstates and the viral titres. From there, the smoothed microstates are calculated by taking the mean over duplicate viral titres, and the result is a null path with the same duplicate titres as the observed data. Repeating this process for each of the 10,000 permutations gives an empirical distribution for the range, which we can then use to calculate a p-value by finding the proportion of random permutations that have a greater test range. The method is further explained in the results section. All GIFT analyses were performed using Python (3.12.8) and numpy (2.1.3), whilst conventional statistics (Kruskal Wallis tests and χ2 tests) were computed using R (4.4.2) and rstatix (0.7.3).

## Results

### Demographic characteristics

The age at sampling was well matched between MS patients and healthy controls, while there was a difference in sex distribution with MS patients more frequently being females keeping in line with the increased female:male ratio observed in MS. EBV seropositivity was significantly higher in MS patients than in controls, validating previous studies. We did not detect differences in seroprevalence of the other herpes viruses between MS patients and healthy controls (Table 1).

**Table 1:** Demographic characteristics of included participants.

|  | **Healthy controls**<br>**(n=137)** | **Multiple sclerosis**<br>**(n=200)** | **p-value** |
| --- | --- | --- | --- |
| **Percentage female\*** | **50%** | **73%** | **$3.9 \times 10^{-5}$** |
| **Age at sampling**<br>**(years, median,**<br>**IQR)** | 38 (27-48) | 39.5 (29.8-47) | 0.93 |
| **Disease duration**<br>**at sampling**<br>**(months, median,**<br>**IQR)** | NA | 8 (3-31.8) | NA |
| **Percentage**<br>**having relapse**<br>**during follow-up** | NA | 15% | NA |
| **Percentage**<br>**having new MRI**<br>**lesions during**<br>**follow up** | NA | 46% | NA |
| **EDSS at sampling**<br>**(median, IQR)** | NA | 1 (0-2) | NA |
| **CMV**<br>**seropositivity\*** | 62.0% | 58.5% | 0.51 |
| **HSV**<br>**seropositivity\*** | 69.3% | 70.5% | 0.82 |
| **VZV**<br>**seropositivity\*** | 97.8% | 97.0% | 0.65 |
| **EBV**<br>**seropositivity\*** | **95.6%** | **100%** | **$2.8 \times 10^{-3}$** |

All comparisons are Mann Whitney U-test for comparison of continuous variables unless indicated otherwise
\* denotes $\chi^2$ with 1 DF
NA: not applicable
EDSS: Expanded Disability Status Scale

### Principle underscoring GIFT on continuous protein data

GIFT is a methodology designed to identify associations between datasets without relying on predefined categories or data grouping and it reduces the dependency of inference precision on sample size. To illustrate the methodology underlying GIFT, consider a specific clinical trait, such as an aggregated continuous measure with three distinct features defined as, f_1_, f_2_ and f_3_, that can be thought of as arising from different disease-related biomarkers. Since the trait values are supposedly linked to the features, assessing potential associations between this continuous trait and the features requires using count or frequency plots, as illustrated in Figs.1A&1B. These plots typically reveal that the different features are observed only within specific ranges of the continuous trait, a pattern confirmed using specific statistical tests comparing the averages noted m_1_, m_2_ and m_3_ in Fig.1C.

**Figure 1:**
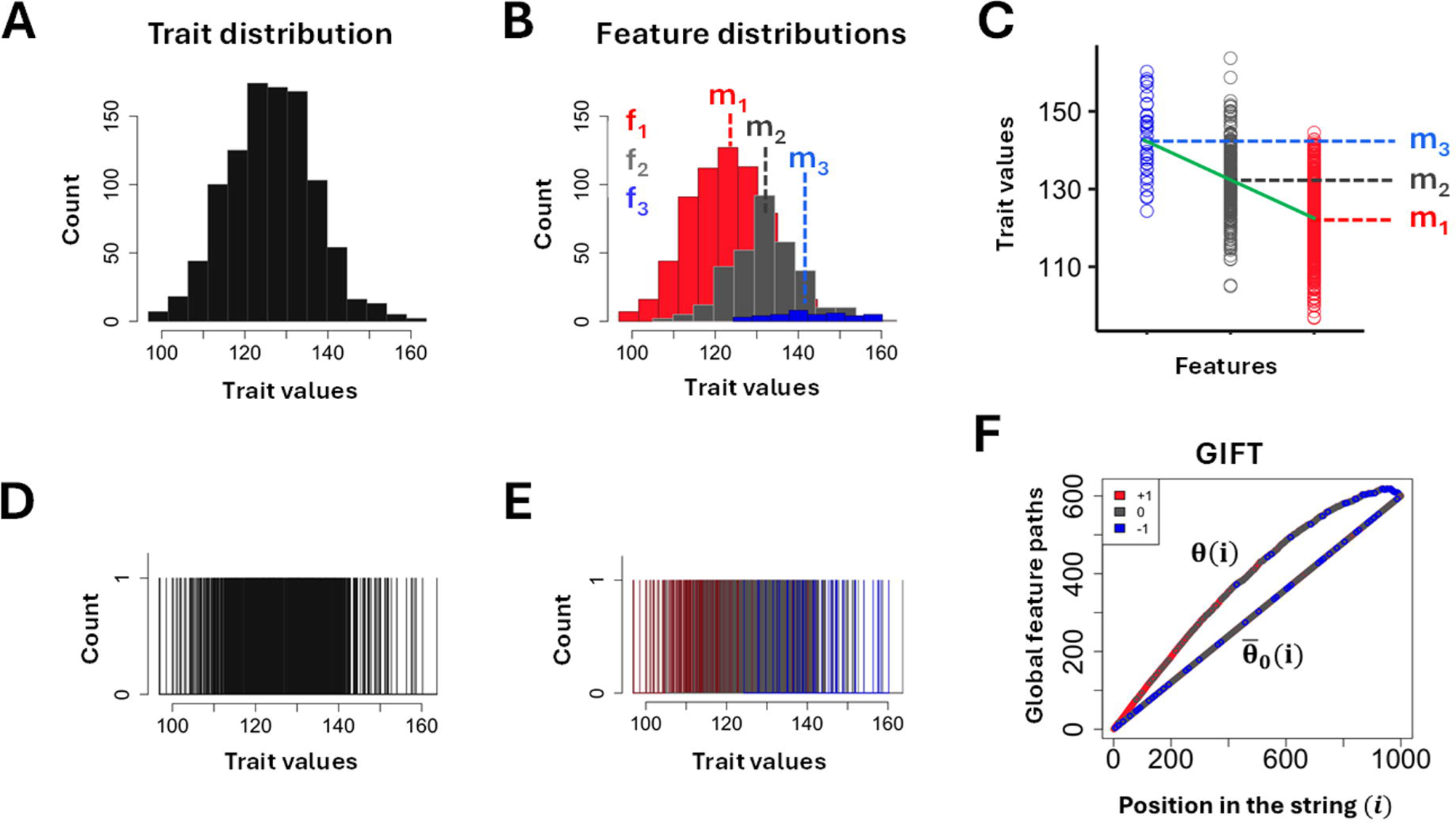
(A) Classic representation of a trait using its distribution density function. (B) If the trait is determined by specific features, each feature is also determined using its distribution density function underlining that different ranges of trait values can be decomposed onto different features. (C) Classically, determining an association between ranges of trait values and features involves determining average differences showing that different ranges of trait values are associated with different features. (D) Contrary to using the distribution density function of the trait, GIFT proceeds by using individual datapoint values for the trait, by way of which the density function is transformed into a barcode. (E) Contrary to using distribution density functions for the features, GIFT proceeds by using individual datapoint values for all features, by way of which the density functions are transformed into a coloured barcode. (F) After numbering the different features, the global cumulative sum of numbered features (*θ*(*i*)) is determined and compared to the resulting average cumulative sum (*θ̅*_0_(*i*)) obtained through an infinite number of permutation of features.

However, this approach may obscure more nuanced associations that GIFT is designed to detect. Indeed, a fundamental limitation of this type of data representation is that substantial information is lost when continuous measures, for instance immunoglobulin levels, are aggregated into categories to provide averages and variances, as variations within a single category cannot be distinguished. GIFT overcomes this limitation by analysing associations without relying on data grouping, preserving the full resolution of the data and enabling precise inferences with smaller sample sizes.

As shown in Figures 1D and 1E, if we were to know the exact value of datapoints, the absence of categories compels us to concentrate on barcodes to provide means to extract inferences. Now, concentrating on the different colours of the bars in Fig.1E, what is visible is that the distribution of colours is not uniform but that bars are segregated as a function of their colour, reflecting the positioning of the different distributions in Fig.1B. Importantly, while each bar is associated with a specific trait value in Fig.1E, this specific trait value depends on the subpopulation (of MS patients) used and a different subpopulation would have provided a slightly different trait value for each bar. That is to say that what matters to determine inferences when considering the coloured barcode in Fig.1E is not the exact positioning of bars, but their relative positioning.

To determine whether the segregation of coloured bars has any significance, GIFT proceeds initially by numbering the different colours using +1 for red bars, 0 for grey bars and –1 for blue bars, thereby creating an initial string of numbers only containing +1, 0 and –1 as they appear from low to high trait values. In a second step, GIFT creates another string where the component, say at position i, corresponds to the cumulative sum of numbers located between the positions 1 to i from the initial string. The component of this new string for the position i is noted *θ*(*i*). Then *θ*(*i*) is plotted as a function of the position i as shown in Fig.1F. The curved aspect of *θ*(*i*) results the presence of red bars (+1 bar) initially followed by grey bars (0 bar) and finally blue bars (-1 bars) as shown in Fig.1E.

This curve is then compared to another function resulting from a scenario corresponding to the null hypothesis. This scenario is obtained by randomly permutating the number +1, 0 and –1 in the initial string. We shall call this randomly permutated string a null string noted *θ*_0_(*i*). However, as there are many random permutations possible of the initial string one may determine the resulting average null string after considering an infinite number of permutations. In this context, it is possible to demonstrate that the resulting average null string leads, after considering the average cumulative sum of components, to a straight-line noted *θ̅*_0_(*i*) (see Fig.1F), whose equation is given by, 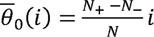, where N_+_, N_-_ are the number of +1, -1 bars, and N is total number of bars in the initial string, respectively. Note that the horizontal bar on the top of 0*_0_* signifies that the average has been considered.

The notion of significance with GIFT is then determined by comparing the difference, *θ*(*i*) - *θ̅*_0_(*i*). There are different ways to determine a measure of significance for the difference *θ*(*i*) - *θ̅*_0_(*i*), which ultimately depend on theoretic considerations such as symmetries and invariances of the dataset and scientific problem studied. However, one generic way to determine a measure of significance is to recall that the segregation of microstates is primordial and that it leads to large amplitudes for *θ*(*i*). One may then concentrate on the difference between the max and min values of, *θ*(*i*) - *θ̅*_0_(*i*). Let us note, Δ, this value. This value (Δ) needs to be compared to a similar value arising when considering the null hypothesis. As *θ̅*_0_(*i*) is an average determined ideally using an infinite number of random permutations, one may for any random permutation *θ*_0_(*i*) realised also determine the difference *θ̅*_0_(i) - *θ*_0_(i) and extract the resulting absolute value of max and min values that we shall note Δ_0_. Repeating the operation a given number of times, fixed to 10,000 times in this paper, then one deduces the probability density, *P*(Δ_0_), that the absolute difference between the max and min values of *θ*_0_(*i*) - *θ̅*_0_(*i*) is Δ_0_. A measure of significance concerning the difference between Δ and Δ_0_ can then be deduced under the form of a p-value using, 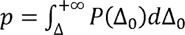.

### MS susceptibility linked to herpes viruses in a sex-dependent manner

Across all viruses, the observed GIFT paths (*θ̅*(*i*)) deviate from the null line (*θ*_0_(*i*)) to varying degrees, indicating differences in the humoral immune response against herpes viruses are linked to MS susceptibility. As expected, higher EBNA1 IgG titres are strongly linked to MS susceptibility in both males and females (Fig. 2), validating previous studies [19]. In addition, also the EBV protein VCA is linked to MS susceptibility across sexes when applying GIFT (Fig. 2A-D), whereas using conventional statistics no association was observed (Table 2). When visually inspecting the GIFT path, most MS patients are in the right part of the GIFT path (indicating high VCA IgG). Importantly, both EBNA1 and VCA are EBV-related proteins, and it is thus not unsurprisingly that both proteins show an association with MS susceptibility. Interestingly, only in males, MS susceptibility is linked to low levels of CMV and HSV IgG (Fig. 2A and C), whereas in females MS susceptibility is linked to high VZV and low HSV IgG (Fig. 2B and D), suggesting a sex-dependent relationships of herpes viruses on MS susceptibility.

**Figure 2:**
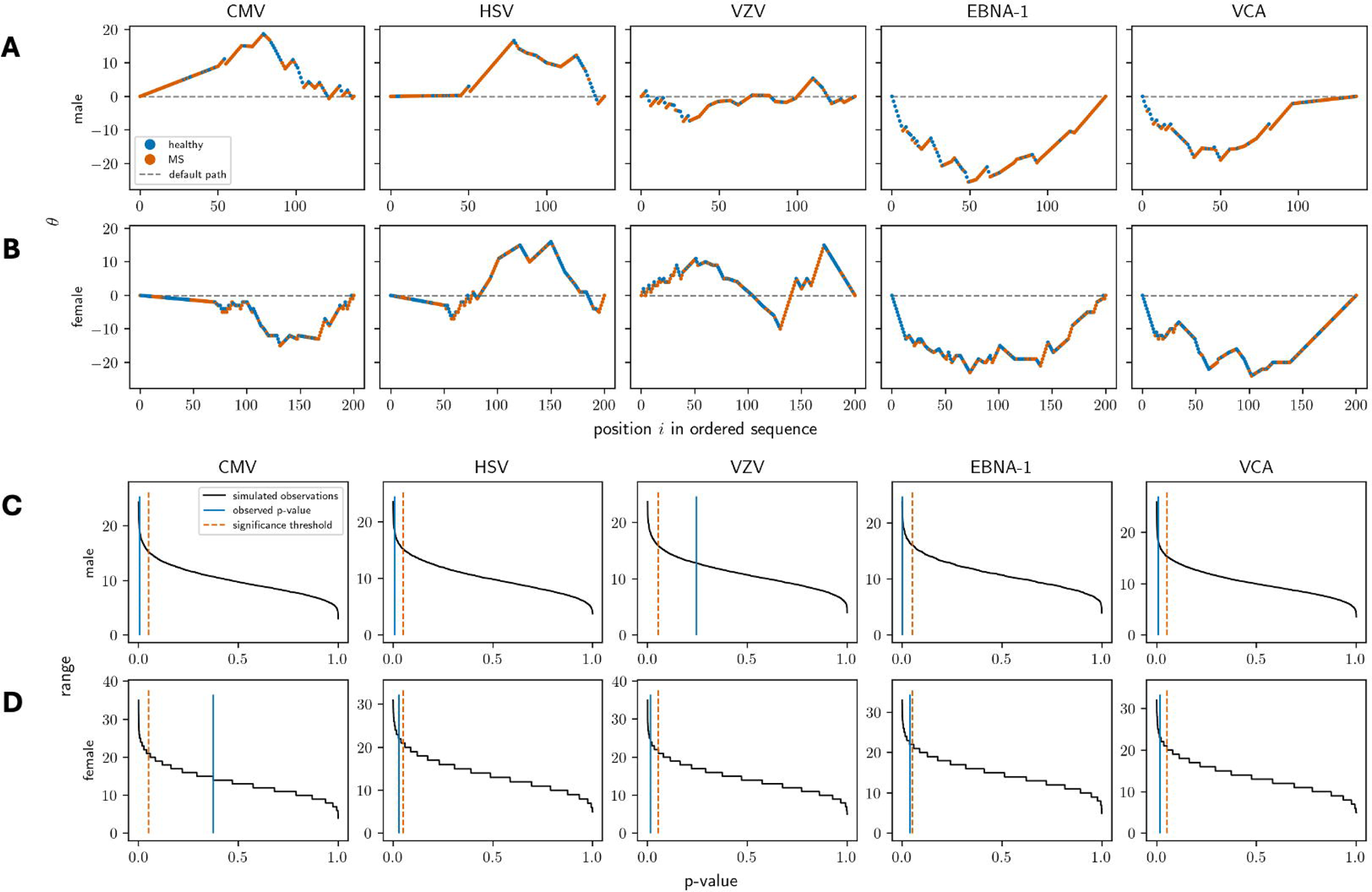
Top panel shows the GIFT paths stratified by gender across for five humoral immune responses against common viral exposures (CMV, HSV, VZV, EBNA1, and VCA). Each subplot portrays the GIFT path obtained using the relevant viral exposure, with A) males displayed in the top row and B) females in the bottom row. The dashed grey lines indicate the null hypothesis. The microstates represent susceptibility for MS (e.g. comparing MS patients with healthy controls). Bottom panel shows the p-values corresponding to the GIFT paths for each virus, stratified according to sex, C) males and D) females. The black lines indicate the range of p-values obtained from simulated null paths for each virus. The blue lines indicate the estimated p-value based on the range in the observed GIFT paths. The orange dashed line indicates the p = 0.05 level.

**Table 2:** humoral immune responses associated with MS susceptibility.

|  | **Females** |  |  | **Males** |  |  |
| --- | --- | --- | --- | --- | --- | --- |
|  | Healthy controls (n=100) | MS (n=100) | p-value | Healthy controls (n=37) | MS (n=100) | p-value |
| **CMV PEI U/ml** | 2001 (2001-2001) | 2001 (2001-2001) | 0.72 | 2001 (2001-2001) | 2001 (2001-2001) | 0.37 |
| **HSV U/ml** | 600 (19-750) | 550 (70.2-700) | 0.37 | **700 (19-1000)** | **380 (19-750)** | **0.046** |
| **VZV mIU/ml** | 1100 (738-1725) | 1100 (588-1525) | 0.32 | 975 (440-1500) | 950 (600-1400) | 0.85 |
| **EBNA1 IgG U/ml** | **25 (14.8-36)** | **35 (23.8-42)** | **$3.4 \times 10^{-4}$** | **18 (5-29)** | **36 (26-42)** | **$2.4 \times 10^{-7}$** |
| **VCA IgG U/ml** | 201 (140-201) | 201 (168-201) | 0.45 | 201 (201-201) | 201 (140-201) | 0.10 |

All values are median and IQR and statistical comparison with Mann Whitney U-test

### Age at sampling is not associated with humoral immune response

Age is a potential confounder, e.g. age at onset in female MS patients is younger than in males, whereas the host immune response against infections is less strong when people are ageing, a phenomena called immunosenescence [20]. Therefore, we next explored the effect age at sampling on the humoral immune response against herpes virus serology. We overlayed the previously obtained GIFT paths with age at sampling and we did not observe a relationship between age and titres for any of the viruses, indicating that the effect of age is negligible (Fig. 3).

**Figure 3:**
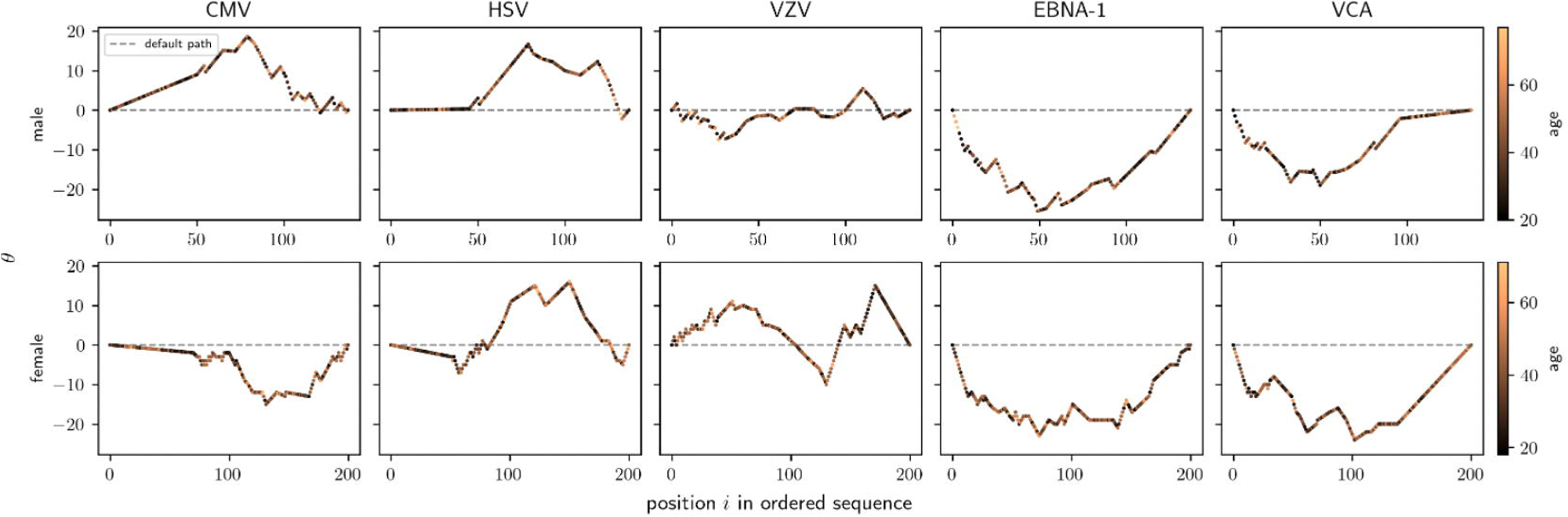
GIFT paths for MS susceptibility are now colour-coded for age at sampling. No skewness of age was observed along the paths.

## Discussion

Modelling high-order data in complex biological traits using conventional statistics has been challenging. In this study, we showed that GIFT can overcome limitation inherit to conventional statistical frameworks. More specifically, we found that the humoral immune response against of some of the common herpes viruses have sex-specific effects associated with MS susceptibility, and that most of those associations remained undetected with conventional statistics.

Because GIFT utilises the full data distribution rather than comparing the averages (e.g. sacrificing important information on exact measurement distribution), it is more powerful to detect subtle and nuanced effects in datasets of complex traits. In addition, GIFT allows visual inspection of the actual data distribution to further understand patterns underlying the data distribution. This potentially allows to detect sensitive cut off points to differentiate groups more accurately, e.g. the exact value where the GIFT path (*θ*(*i*)) changes from a negative to a positive slope or vice versa.

EBV has consistently been associated with MS susceptibility [19] and several biological mechanisms have been shown how EBV contributes to a person’s risk to develop MS after EBV infection [21]. In a recent meta-analysis, also HHV-6 and VZV were found to be associated with MS susceptibility [22]. In our study, VZV was not significant with conventional statistics, however GIFT identified an association with female susceptibility for MS. Conflicting results on MS susceptibility and CMV have been published, which is at least partially attributed to small sample size of studies and potential regional or ethnic differences [23]. Systematic comparisons of immunoglobulin levels against herpes viruses between sexes have not been performed yet, likely due relatively limited sample sizes and lack of statistical power using conventional statistical frameworks. GIFT has the ability to overcome this limitation as it is more powerful to detect associations in small datasets [14]. Therefore, our results are of interest as it highlights a sex-dependent effect of the humoral immune response against common viruses and MS susceptibility.

Our study has some limitations. Although GIFT is more powerful than conventional statistics to work with small datasets, the number of patients and controls in the current study is relatively limited. Because of this, we could not divide the total dataset in a training and validation cohort. Moreover, our study lacks a replication cohort. Finally, we have only included participants from Northern Italy, and we cannot generalise our findings immediately to other geographical areas or non-White European populations. Therefore, independent replication of our results in ethnic diverse populations will be necessary.

Taken together, we have shown for the first time that GIFT is reliable tool to detect subtle associations in human protein datasets, and that those associations remained undetected using conventional statistical frameworks.

## Data Availability

All data produced in the present study are available upon reasonable request to the authors and upon completion of a signed data transfer agreement

## Funding

This research was funded in part by The Italian Multiple Sclerosis Foundation (FISM), grant number 2020 R-Multi 050, to B.G. R.T. has received support from the UK MRC (CARP MR/T024402/1).

## Conflict of interest

None of the authors report a conflict of interest

## Notes

### Competing Interest Statement

The authors have declared no competing interest.

